# Dexmedetomidine decreases cerebral hyperperfusion incidence following carotid stenting: A randomized, double-blind trial

**DOI:** 10.1101/2023.07.06.23292333

**Authors:** Enqiang Chang, Lingzhi Wu, Xinyi Li, Jinpeng Zhou, Hui Zhi, Min Sun, Guanyu Chen, Li Li, Tianxiao Li, Daqing Ma, Jiaqiang Zhang

## Abstract

**Background:** Cerebral hyperperfusion syndrome (CHS) is not a common but severe complication after carotid artery stenting (CAS). We investigated whether prophylactic low-dose dexmedetomidine, a selective α_2_-adrenoceptor agonist, can decrease cerebral hyperperfusion induced brain injury after carotid artery stenting.

**Methods:** This randomised, double-blind, placebo-controlled trial was conducted in a tertiary-care hospital in Zhengzhou, China. Patients aged 18 to 80 years old who had undergone CAS and from whom written informed consent was obtained, were enrolled between Jul 20, 2019 and Oct 10, 2022. Patients were randomly assigned to receive either intravenous dexmedetomidine (0·1 μg/kg/h, from insertion of the laryngeal mask until 72hr on postoperative day 3) (n = 80) or placebo (intravenous normal saline) (n = 80). The primary endpoint was the incidence of cerebral hyperperfusion (CH) and cerebral hyperperfusion syndrome (CHS), which were assessed six times with Transcranial Doppler sonography (TCD) during the postoperative three days. This study was registered with the Chinese Clinical Trial Registry, www.chictr.org.cn, ChiCTR1900024416.

**Results:** CH occurred in 30 (37·5%) of 80 patients given a placebo and in 9 (11·2%) of 80 patients given dexmedetomidine (odds ratio [OR] 0·211, 95% CI 0·09-0·48; p<0·001). Further, CHS was significantly lower in the Dex group than in the placebo group (2.5% vs 13·75%; [OR] 0·16, 95% CI 0·03-0·71; p=0·017). Correspondingly, dexmedetomidine significantly upregulated serum brain-derived neurotrophic factor (BDNF) and downregulated neuronal injury biomarker neurofilament light chain (Nfl). The hierarchical clustering analysis revealed little difference in lipids metabolites between the two groups pre- and post-operatively, with Dex treatment uniquely increased lysphosphatidylethanolamine (LPE).

**Conclusions:** A low prophylactic dose of dexmedetomidine significantly decreased the occurrence of cerebral hyperperfusion and cerebral hyperperfusion syndrome during the first three days after carotid artery stenting surgery.

## INTRODUCTION

Stroke remains the second-leading cause of death and the third-leading cause of disability worldwide. It has been estimated that one-third of all ischemic strokes result from carotid artery stenosis and plaque shedding.^1^ Carotid artery stenting (CAS) is increasingly used as the primary treatment for carotid artery stenosis due to its minimal trauma, rapid recovery, and effectiveness in preventing stroke recurrence.^2^

Due to chronic cerebral vascular stenosis, intracerebral arteries are highly dilated, and cerebrovascular reserve capacity is compromised. When cerebral blood flow (CBF) is resumed in patients following surgery, ipsilateral blood vessels fail to constrict promptly and sufficiently. As a result, there is a sudden increase in the volume and blood flow into the ipsilateral cerebral hemisphere to become hyperperfused. In addition, extensive leakage of plasma components into the cerebral tissues due to minor vascular injuries results in cerebral vasogenic edema and intracranial hypertension.^3^ Consequently, a less common but severe complication, cerebral hyperperfusion syndrome (CHS) occurs. CHS after CAS has an estimated occurrence of up to 40%; untreated individuals eventually develop hemorrhagic stroke or even die.^4^ Currently, there is no effective treatment available once CHS occurred. Therefore, the current strategy is to prevent CHS during and after stenting surgery through vigilant monitoring and careful management. Titrating cerebral blood flow to avoid excessive perfusion and minimizing postoperative hemorrhage risk is part of systematic approaches for patients’ better outcomes.

Dexmedetomidine (Dex) is an α_2_-adrenoceptor agonist with sedative, anxiolytic, sympatholytic, and analgesic-sparing effects.^5^ Furthermore, Dex was shown to reduce cerebral blood flow (CBF) velocity in dogs without affecting cerebral metabolic rate^6^. Dex also was reported to have potent neuroprotective properties.^7^ Indeed, it has been used in patients with traumatic brain injury and cerebral edema, and the outcomes are promising.^8^ Furthermore, sleep disturbances are a common problem after stenting due to various reasons, including headaches or seizures, focal neurological deficits, and visual disturbances.^9^ Low-dose dexmedetomidine infusion at 0·1 μg/kg/h was reported to be safe and significantly improve sleep quality in surgical and critically ill patients. ^5^. In view of the foregoing, it is plausible for Dex to decrease the risk of CH and CHS in patients after CAS. We, therefore, designed the current randomized controlled trial to investigate whether a prophylactic intravenous infusion of dexmedetomidine prevents CH and CHS in patients after CAS with blood lipidomics analyses, neuronal injurious and protective biomarker measurement, and clinical outcome assessments.

## MATERIAL AND METHODS

### Ethics statement

The study protocol was approved by the Ethics Committee of People’s Hospital of Zhengzhou University (2019-43), China, and registered with the Chinese Clinical Trial Registry (www.chictr.org.cn; ChiCTR1900024416). The protocol and study procedures comply with the Regulations on Administration of Human Genetic Resources, administered by the Ministry of Health, Beijing, China. In addition, written informed consents were obtained from all participating patients who were scheduled to recruit after screening assessments or from their next of kin or their legal representative.

### Patient’s recruitments

This randomized, double-blinded, parallel-arm placebo-controlled trial was conducted between July 2019 and December 2022 at the Zhengzhou University People’s Hospital, China. The primary aim of this study was to evaluate the superiority of the intervention (dexmedetomidine). The inclusion criteria included patients aged 18 to 80 who underwent elective CAS under general anesthesia (GA)^10^. Patients were excluded if they met any of the following criteria: 1· inability to communicate in the preoperative period (coma, profound dementia, or language barrier), 2· non-intracranial atherosclerotic stenosis (ICAS), 3· acute ischemic stroke within two weeks, 4· cardiogenic embolism, 5· where TCD couldn’t be performed (no temporal window or anatomical malformations), mRS>3, NIHSS >6, 6· unsatisfactory surgical effect (residual stenosis rate > 30%, detachment of thrombus or CEA failure), 7· known preoperative sick sinus syndrome, severe sinus bradycardia (<50 beats per min [bpm]), or second degree or greater atrioventricular block without a pacemaker, and 8· patients refused to participate. All patients were required to fast for 8 hours before surgery.

### Randomization and masking

SAA9.4 (SAS Institute, Cary, NC) was used to generate random numbers with a 1:1 ratio, done by a biostatistician independent for data collection, management, and statistical analysis. Recruited patients were randomly assigned to the Dex group (dexmedetomidine) or the placebo group (normal saline). The trial pharmacist (unblinded/ independent) worked as an administrator to manage and distribute the study drugs based on randomization. The randomization process ensured concealed allocation and blinding of the specialist, the participant, and the outcome assessor during recruitment, data collection, and analysis during the study period.

### Procedures

#### Carotid angioplasty and stenting (CAS)

Patients’ baseline data and blood samples of patients were collected before the operation, and continuous invasive radial arterial blood pressure monitoring was established under local anesthesia. Anesthesia was induced with midazolam, etomidate, sufentanil, and rocuronium and maintained with propofol infusion with our routine practice. The laryngeal mask was then inserted, and their lungs were ventilated. The intraoperative mean arterial pressure (MAP) was strictly maintained with ≤ ±10 mmHg increase from the preoperative level until the carotid stenting was opened. If necessary, intravenous norepinephrine was infused continuously at 2 to 6 μg/kg/h to maintain optimal MAP. After completing the CAS, urapidil maintained MAP between 70-105 mmHg.^2^ Patients were extubated when they met the criteria and stayed in the post-anesthesia care unit (PACU) for at least 6 h.

#### Study drugs

Dexmedetomidine hydrochloride in 200μg/2mL and 2mL normal saline were provided as clear aqueous solutions in identical 3 mL bottles (manufactured by Jiangsu Hengrui Medicine Co, Ltd, Jiangsu, China). Before administration, they were diluted with normal saline to 200 mL, and the final concentration of dexmedetomidine hydrochloride was 1 μg/mL. Continuous dexmedetomidine infusion was started after anesthesia induction at a rate of 0·1 μg /kg/hr for 72h.

#### Transcranial Doppler sonography (TCD) measurement

Middle cerebral artery blood velocity (MCAV)^11^ was determined at the following points: pre-, intra-(before stenting and 1 min after stenting), and post-operatively (at 6, 24, 48, 72 h) with Pulse Doppler transducer (EMS-9PB, Delica Medical, Shenzhen, China) with a 1·6-MHz probe. In order to access the MCA ipsilateral to the operated carotid artery, the transducer was adjusted in a head frame, and the values were collected in real time for further offline analysis. TCD measurements were performed at seven pre-defined peri-operative moments when the MAP was within the above range (Figure S1). The mean velocity (*Vmean*) in the MCA ipsilateral to the treated carotid artery was measured: The intra-operative *Vmean* change ((*Vpost-stent* – *Vpre-stent*)/*Vpre-stent*) was compared with the post-operative data at postoperative 6, 24, 48 and 72h ((*Vpost-op6, 24, 48* and *72h*-*Vpre-op*)/*Vpre-op*) in relation to CHS (Figure S1).^12^

### Study endpoints

The primary endpoint was the incidence of CH and CHS in the first three days after the operation. CH was defined as the increase of TCD-derived MCAV_mean_ ≥ 100% compared with the pre-operative baseline MCAV_mean_. CHS was defined as CH combined with clinical symptoms such as confusion, seizures, and focal neurological deficits after a symptom-free interval or imaging examination that revealed intracranial edema and/or intracranial hemorrhage; headache, usually on the same side of the surgery; sharp, abrupt in postoperative blood pressure to the level of malignant hypertension [systolic blood pressure >180 mmHg, diastolic blood pressure > 100 mmHg]. Clinical diagnosis of CHS was made by 2-3 independent neurologists if any of the above indications were met.^13^ Outcome assessment was conducted by trained researchers who did not participate in patient clinical care. Secondary endpoints included NIHSS and mRS within 30 days after the operation, extubation time, discharging from the hospital within seven days post-operation, length of stay in hospital post-operation, and all-cause 30-day mortality.

#### Other outcomes

postoperative pain intensity and subjective sleep quality were assessed with the Numeric Rating Scale (NRS). Pain and sleep assessments were performed only if the Richmond Agitation Sedation Scale (RASS)^14^ score was − 2 to +1. Exploratory analyses included the incidence of CH according to the duration of the TCD test, time to onset and duration of CHS, and the difference in laboratory findings before and after the operation were compared.

### Adverse event treatment

Healthcare personnel strictly monitor adverse events until the event resolves or 72 hours after CAS. Patients with a heart rate below 55 beats/min or a decrease of more than 20% from baseline are defined as bradycardia (in the case of baseline values [before entry into the study drug] ≤ 69 beats/min). Systolic blood pressure decreased by more than 20% from baseline when systolic blood pressure was less than 95 mmHg, or a baseline value of less than 119 mmHg is defined as hypotension. A heart rate > 100 beats/min or an increase of more than 20% from baseline at a baseline of > 83 beats/min is defined as tachycardia. Hypertension is defined as a systolic blood pressure > 160 mmHg, or an increase in systolic blood pressure by more than 20% from baseline at a baseline of 133 mmHg. Pulse oximetry below 90% of the baseline is defined as hypoxemia. Interventions for bradycardia, tachycardia, and hypertension included adjusting the infusion or administration of study drugs, or both. Interventions for hypotension included adjustment of study drug infusions, intravenous fluids, or administration of both. Interventions for hypoxemia include oxygen therapy (for patients not intubated), adjustment of ventilator settings (for intubated patients), or physiotherapy.

### Lipidomics

Ten patients from each group were randomly chosen to collect blood samples (10 ml/time point) before anaesthesia induction and 6h post-operation. Ethylenediaminetetraacetic sodium acid (EDTA) was used to coat the tubes to prevent clotting for plasma separation with centrifugation at 2000 x g for 10 minutes at 4°C Plasma samples were stored at -80°C unless metabolite and lipid analysis. For metabolite extraction, a 100μl sample was mixed with 400 μL of methanol containing internal standards for analysis. The mixture was vortexed for 2 mins and then centrifuged at 15,000 x g for 15 mins at 4 °C. Supernatant was collected with approximately 200μl per sample for LC-MS analysis. For lipid extraction, 300 μl of methanol containing internal standards was mixed with 50 μl of serum sample. The mixture was vortexed for 3 minutes, and then 1000 μl of MTBE and 200 μl of ultrapure water were added. Afterwards, the mixed samples were shaken at 37°C for 15 minutes and left at 4°C for 30 minutes for separation. The upper lipid layer (900 μl) was transferred and dissolved in 600 μl of an acetonitrile-isopropanol mixture for further non-targeted lipidomics analysis with the QEXactive mass spectrometry, and LipidSearch™ software (OPTON-30880, version 5·0) was used for lipid identification and data process.

### Brain-derived neurotrophic (BDNF) and Neurofilament Light (Nfl) measurement

Internal jugular vein blood was collected into a serum separator tube before anaesthesia induction and at postoperative 24h and 72h in the medium care unit (MCU). Samples were centrifuged at 2,000 x g for 10 minutes after clot formation to collect serum. Serum NfL concentrations were measured in duplicates according to the manufacturer’s instructions on a Single Molecule Array Measurement HD-X Analyzer platform (Quanterix, USA) using the NF-Light Advantage Kit (103186, Quanterix). In addition, serum BDNF concentrations were measured with the human BDNF Elisa kit (ab212166, Abcam, UK).

### Statistical analysis

#### Clinical data

Our preliminary study showed that the incidence of cerebral hyperperfusion in the Placebo and the Dex group was 36·7% (11/30) and 13·3% (4/30), respectively. With α=0·05 and β=0·1 using Fisher’s exact test to determine the difference of cerebral hyperperfusion incidence between the two groups, we determined that the minimum sample size would need to be 140 cases. Considering potential patients and data loss during the study, the sample size was inflated by 10%, and the total number of patients was 160 (80/arm).

The patients’ characteristics were analyzed with the Fisher’s exact test (categorical variables), independent t-test, or Mann-Whitney U test (continuous variables) as appropriate for comparing two groups. The Hodges-Lehmann estimator calculated the difference between the two medians with 95% CI. The Kaplan-Meier estimator was used to estimate the time-to-event results, and the log-rank test was used to analyze the differences between groups. SPSS version 14·0 (SPSS, Chicago, IL) and SAS version 9·2 software (SAS Institute, Cary, NC) were used to process the statistical analysis with two-tailed tests as appropriate. The significance was considered when the p-value was less than 0·05. The Clinical Research Ethics Committee from Zhengzhou University People’s Hospital oversaw the data throughout the study.

#### Lipidomics

Multivariate statistical analysis was performed in this part, and the unsupervised principal component analysis (PCA) was used to evaluate the population distribution and stability between samples during the process. The supervised partial least squares (PLS-DA) and orthogonal partial least squares analysis (OPLS-DA) were used to analyse the difference in the lipidomics between the two groups. Student t-test and fold change analysis were used to compare differential metabolites between the two groups. A P-value less than 0·05 was considered statistically significant. The volcano map was used to visualise the P values from the t-test and fold changes from the fold change analysis. All significant differential metabolites were further analysed with Hierarchical Clustering analysis. The heat map visualised the differential metabolites of the top 50 of the most significant. Volcano maps were used to visualise differential metabolites’ P, VIP and Fold change values.

## RESULTS

### Patients’ characteristics

Between Jul 20, 2019 and Oct 10, 2022, 381 patients were screened, and 221 patients were excluded (including 189 patients who met the exclusion criteria and 32 patients who refused to participate). The remaining 160 patients were enrolled in the trial and randomly assigned to either the dexmedetomidine or placebo group (n=80, respectively). There were no lapses during the study. The study drug infusion was modified for 14 patients due to adverse events. Enrollment was stopped upon attainment of recruitment goals (Figure 1). The final visit of the last randomized patient was on Nov 10, 2022.

**Figure 1.**
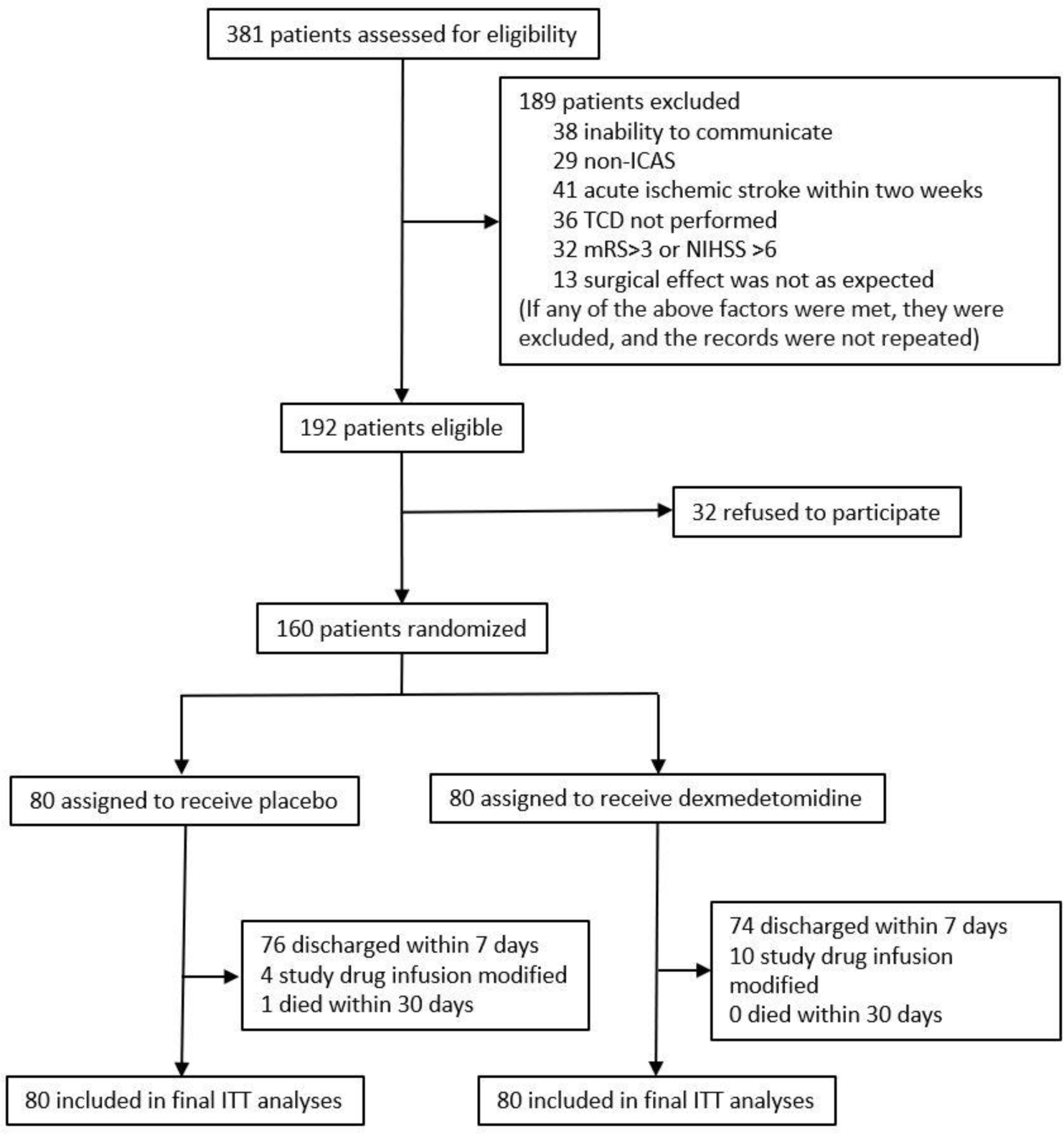
Trial profile. ITT analyses included all randomized patients in the groups to which they were randomly assigned. ITT=intention-to-treat. ICAS: intracranial atherosclerotic stenosis. TCD: Transcranial doppler. mRS: Modified Rankin. NIHSS: National Institute of Health stroke scale.

The patient’s baseline characteristics and perioperative variables related to surgery and anesthesia are listed in Table 1. Overall, both groups were well-matched for baseline and perioperative variables. The proportion of patients receiving additional sedation after randomization was similar in both groups. The total dose of propofol used for sedation was lower in the Dexmedetomidine group than in the placebo group (Table 1).

**Table 1.** Perioperative variables.

| Variables | Placebo group (n=80) | Dex group (n=80) |
| --- | --- | --- |
| **Preoperative** |  |  |
| Gender (male) | 53 (66.2%) | 56 (70.0%) |
| Age (y) | 65.4 (10.46) | 66.2 (9.84) |
| BMI (score) | 24.8 (3.76) | 25.3 (3.66) |
| Hypertension | 57 (71.2%) | 53 (66.2%) |
| Hypercholesterolemia | 37 (46.2%) | 42 (52.5%) |
| Diabetes mellitus | 19 (23.8%) | 17 (21.2%) |
| Smoking | 23 (28.8%) | 26 (32.5%) |
| Alcohol use | 29 (36.2%) | 33 (41.2%) |
| MAP (mmHg) | 108.6 (17.4) | 106.5 (18.4) |
| Symptomatic (TIA, Cerebral infarction) | 65 (81.3%) | 68 (85%) |
| Operation side (right) | 41 (51.2%) | 37 (46.2%) |
| Degree of ipsilateral stenosis |  |  |
| 50%-70% | 12 (15%) | 14 (17.5%) |
| >70% | 68 (85%) | 66 (82.5%) |
| Degree of contralateral stenosis |  |  |
| <50% | 36 (45%) | 38 (47.5%) |
| 50%-70% | 30 (37.5%) | 28 (35%) |
| >70% | 11 (13.8%) | 13 (16.2%) |
| Occlusion | 3 (3.75%) | 1 (1.25%) |
| Modified Rankin (mRS) (score) | 1 (0-3) | 1 (0-3) |
| NIHSS (score) | 3 (1-5) | 3 (1-5) |
| Preoperative WBC count (x10<sup>3</sup>/μl) | 6.62 (1.47) | 6.34 (1.39) |
| Preoperative C-reactive protein (mg/L) | 5.7 (3.1) | 5.5 (3.6) |
| **Intra- and Post-operative** |  |  |
| Duration of operation (min) | 95 (62-130) | 90 (56-135) |
| Duration of general anesthesia (min) | 125 (90-160) | 120 (85-165) |
| Intraoperative fluid balance (ml) | 833 (450-1200) | 861 (460-1400) |
| MAP (mmHg) | 87.1 (15.1) | 85.9 (14.7) |
| PaCO<sub>2</sub> (mmHg) | 35 (31-37) | 36 (33-38) |
| Residual stenosis rate |  |  |
| 0-10% | 10 (12.5%) | 11 (13.8%) |
| 10%-20% | 38 (47.5%) | 41 (51.3%) |
| 20%-30% | 32 (40.0%) | 28 (35.0%) |
| Medicine |  |  |
| Dexamethasone* | 73 (91.3%) | 69 (86.2%) |
| Atropine | 6 (7.5%) | 10 (12.5%) |
| Norepinephrine | 36 (45.0%) | 33 (41.3%) |
| General anesthetics |  |  |
| Propofol (mg) | 513 (82.1) | 426 (87.8) |
| Midazolam (mg) | 3.75 (0.935) | 3.66 (0.871) |
| Sufentanil (µg) | 15.8 (6.19) | 15.3 (6.88) |

\*For prophylaxis of postoperative nausea and vomiting, data are expressed as number (%), mean (SD) or median (interquartile range). Dex: dexmedetomidine; BMI= Body Mass Index, MAP= mean arterial pressure, NIHSS= National Institute of Health stroke scale, mRS= Modified Rankin, MCAV mean= mean blood flow velocity in the middle cerebral artery.

Cerebral hyperperfusion occurred in 30 (37·5%) of 80 patients in the Placebo group and 9 (11·2%) of 80 patients in the Dexmedetomidine group (odds ratio [OR] 0·211, 95% CI 0·09-0·48; p<0·001). Cerebral hyperperfusion was most frequently reported on stent opening and at 48h postoperative in both groups (Figure S2). Further, cerebral hyperperfusion syndrome was significantly lower in the Dex group than in the placebo group (2·5% *vs* 13·75%; [OR] 0·16, 95% CI 0·03-0·71; p=0·017) (Table 2).

**Table 2.**
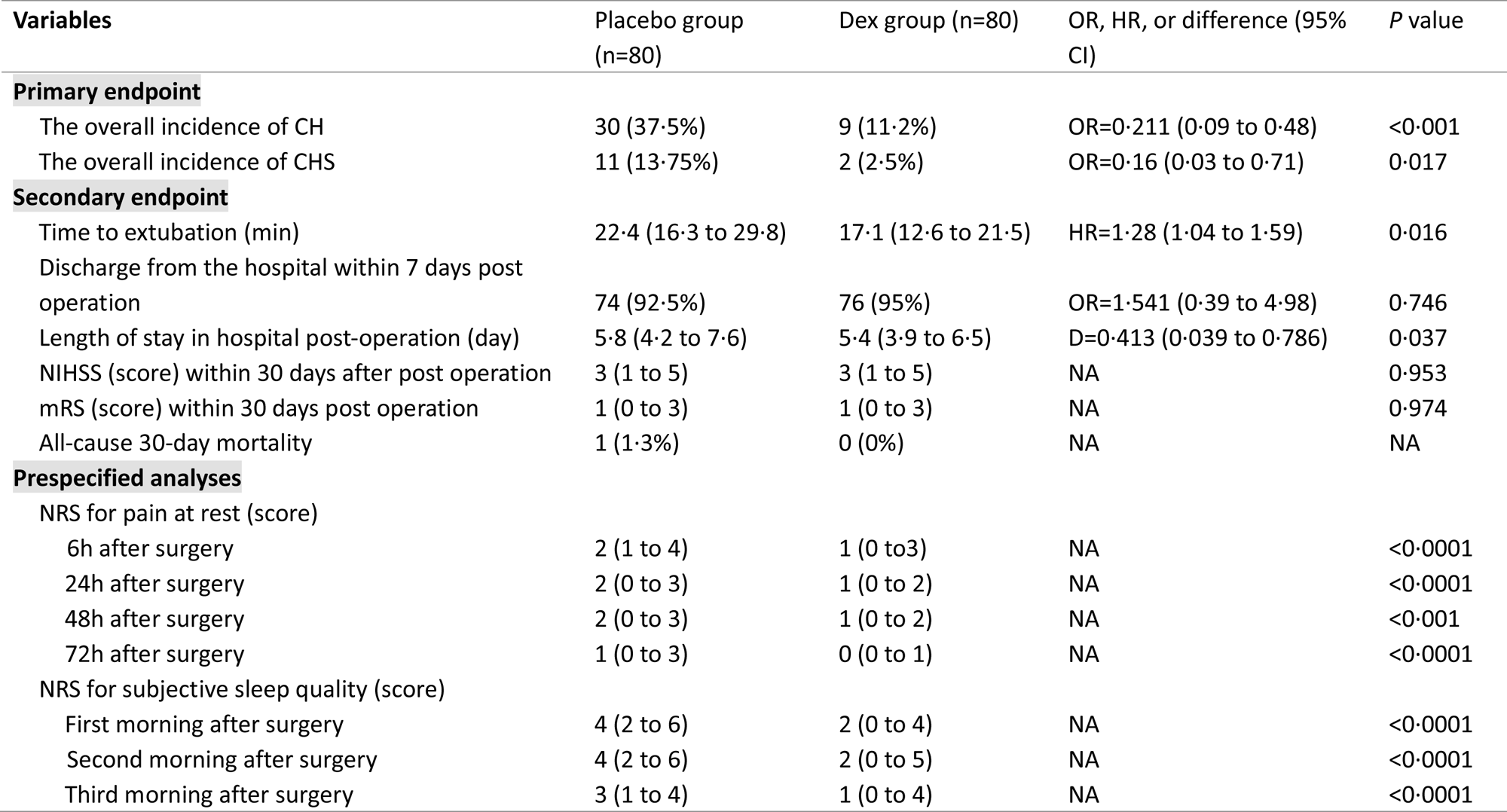
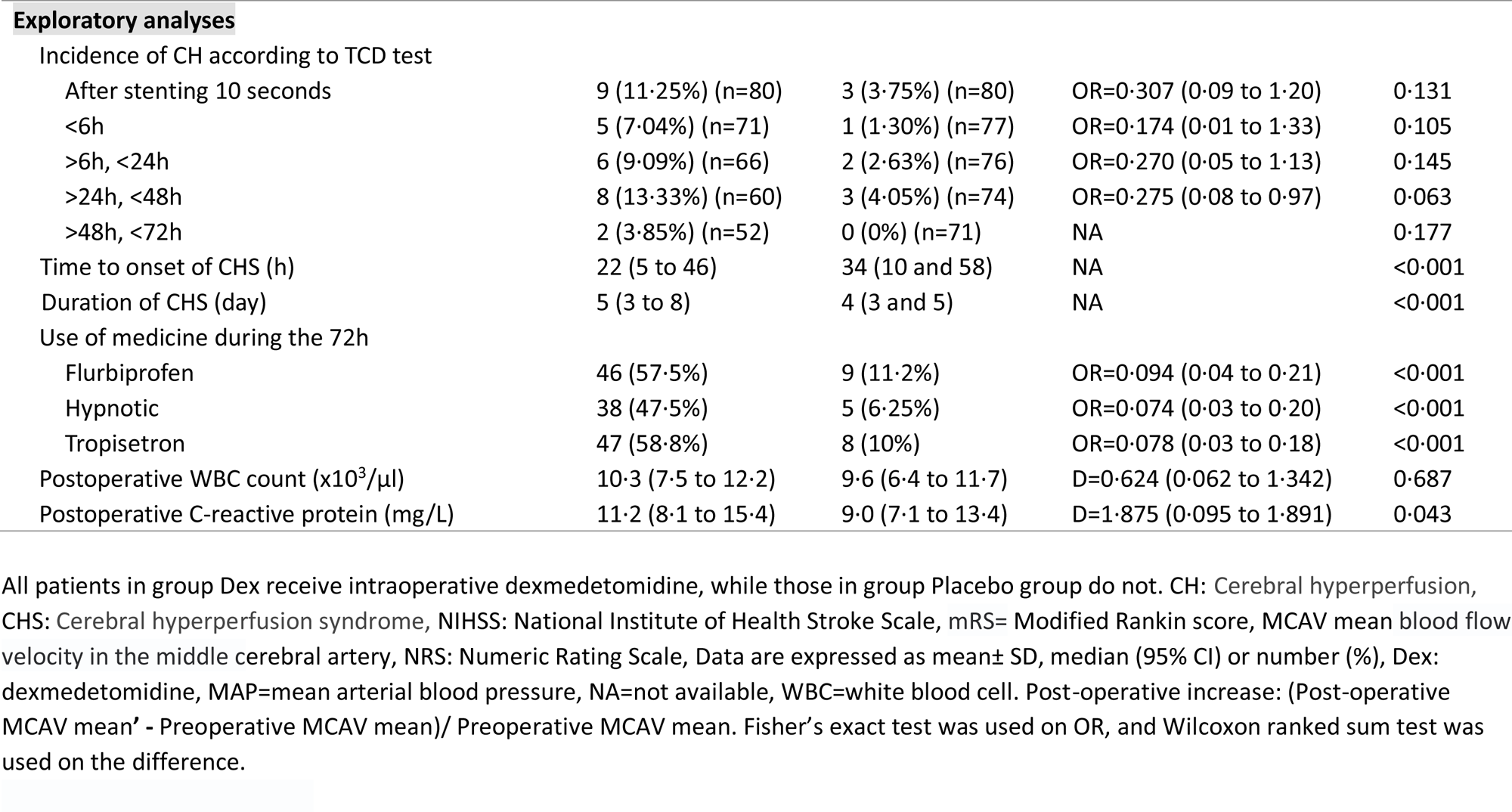
Effectiveness outcomes data.

| **Table 2 Effectiveness outcomes data** |  |  |  |  |
| --- | --- | --- | --- | --- |
| **Variables** | **Placebo group**<br>**(n=80)** | **Dex group (n=80)** | **OR, HR, or difference (95%**<br>**CI)** | **P value** |
| **Primary endpoint** |  |  |  |  |
| The overall incidence of CH | 30 (37.5%) | 9 (11.2%) | OR=0.211 (0.09 to 0.48) | <0.001 |
| The overall incidence of CHS | 11 (13.75%) | 2 (2.5%) | OR=0.16 (0.03 to 0.71) | 0.017 |
| **Secondary endpoint** |  |  |  |  |
| Time to extubation (min) | 22.4 (16.3 to 29.8) | 17.1 (12.6 to 21.5) | HR=1.28 (1.04 to 1.59) | 0.016 |
| Discharge from the hospital within 7 days post<br>operation | 74 (92.5%) | 76 (95%) | OR=1.541 (0.39 to 4.98) | 0.746 |
| Length of stay in hospital post-operation (day) | 5.8 (4.2 to 7.6) | 5.4 (3.9 to 6.5) | D=0.413 (0.039 to 0.786) | 0.037 |
| NIHSS (score) within 30 days after post operation | 3 (1 to 5) | 3 (1 to 5) | NA | 0.953 |
| mRS (score) within 30 days post operation | 1 (0 to 3) | 1 (0 to 3) | NA | 0.974 |
| All-cause 30-day mortality | 1 (1.3%) | 0 (0%) | NA | NA |
| **Prespecified analyses** |  |  |  |  |
| NRS for pain at rest (score) |  |  |  |  |
| 6h after surgery | 2 (1 to 4) | 1 (0 to 3) | NA | <0.0001 |
| 24h after surgery | 2 (0 to 3) | 1 (0 to 2) | NA | <0.0001 |
| 48h after surgery | 2 (0 to 3) | 1 (0 to 2) | NA | <0.001 |
| 72h after surgery | 1 (0 to 3) | 0 (0 to 1) | NA | <0.0001 |
| NRS for subjective sleep quality (score) |  |  |  |  |
| First morning after surgery | 4 (2 to 6) | 2 (0 to 4) | NA | <0.0001 |
| Second morning after surgery | 4 (2 to 6) | 2 (0 to 5) | NA | <0.0001 |
| Third morning after surgery | 3 (1 to 4) | 1 (0 to 4) | NA | <0.0001 |
| Exploratory analyses |  |  |  |  |
| Incidence of CH according to TCD test |  |  |  |  |
| After stenting 10 seconds | 9 (11.25%) (n=80) | 3 (3.75%) (n=80) | OR=0.307 (0.09 to 1.20) | 0.131 |
| <6h | 5 (7.04%) (n=71) | 1 (1.30%) (n=77) | OR=0.174 (0.01 to 1.33) | 0.105 |
| >6h, <24h | 6 (9.09%) (n=66) | 2 (2.63%) (n=76) | OR=0.270 (0.05 to 1.13) | 0.145 |
| >24h, <48h | 8 (13.33%) (n=60) | 3 (4.05%) (n=74) | OR=0.275 (0.08 to 0.97) | 0.063 |
| >48h, <72h | 2 (3.85%) (n=52) | 0 (0%) (n=71) | NA | 0.177 |
| Time to onset of CHS (h) | 22 (5 to 46) | 34 (10 and 58) | NA | <0.001 |
| Duration of CHS (day) | 5 (3 to 8) | 4 (3 and 5) | NA | <0.001 |
| Use of medicine during the 72h |  |  |  |  |
| Flurbiprofen | 46 (57.5%) | 9 (11.2%) | OR=0.094 (0.04 to 0.21) | <0.001 |
| Hypnotic | 38 (47.5%) | 5 (6.25%) | OR=0.074 (0.03 to 0.20) | <0.001 |
| Tropisetron | 47 (58.8%) | 8 (10%) | OR=0.078 (0.03 to 0.18) | <0.001 |
| Postoperative WBC count (x10<sup>3</sup>/μl) | 10.3 (7.5 to 12.2) | 9.6 (6.4 to 11.7) | D=0.624 (0.062 to 1.342) | 0.687 |
| Postoperative C-reactive protein (mg/L) | 11.2 (8.1 to 15.4) | 9.0 (7.1 to 13.4) | D=1.875 (0.095 to 1.891) | 0.043 |

All patients in group Dex receive intraoperative dexmedetomidine, while those in group Placebo group do not. CH: Cerebral hyperperfusion, CHS: Cerebral hyperperfusion syndrome, NIHSS: National Institute of Health Stroke Scale, mRS= Modified Rankin score, MCAV mean blood flow velocity in the middle cerebral artery, NRS: Numeric Rating Scale, Data are expressed as mean± SD, median (95% CI) or number (%), Dex: dexmedetomidine, MAP=mean arterial blood pressure, NA=not available, WBC=white blood cell. Post-operative increase: (Post-operative MCAV mean' - Preoperative MCAV mean)/ Preoperative MCAV mean. Fisher's exact test was used on OR, and Wilcoxon ranked sum test was used on the difference.

After surgery, the extubation time was longer in the Placebo group than in the Dexmedetomidine group (22·4 min [95% CI 16·3–29·8] vs 17·1 min [12·6–21·5] [hazard ratio (HR) 1·28, 95% CI 1·04–1·59; p=0·016]). No significant difference in hospital discharge within seven days post-operation was found between the two groups; However, the post-analysis revealed that the length of stay in hospital after the CAS was shorter in the Dexmedetomidine group (5·8 d [4·2 to 7·6]) than in the placebo group (5·4 d [3·9 to 6·5]), p=0·037. At 30 days post-operation, NIHSS, mRS and all-cause mortality were not significantly different between the two groups (table 2). A case in the Placebo group suddenly developed a coma on day five after surgery; CT showed extensive hemorrhage in the ipsilateral cerebral hemisphere and died (Figure S3).

The NRS pain scores in the Dex group were significantly lower than in the placebo group after surgery up to 72h (all p-values < 0·001). In addition, the subjective sleep quality scores were significantly lower in the Dex group than that of the placebo group up to post-operation day 3 (All p-values <0·0001; Table 2). For the exploratory analyses, the incidence of cerebral hyperperfusion at each TCD timeframe was not different between the two groups; however, the time to onset and duration of CHS differed. Furthermore, medication and C-reactive protein were significantly lower in the Dex group than in the Placebo group during the postoperative period (table 2).

The RASS scores were no different between the Dex and placebo groups. There was no significant difference in TIA, cerebral infarction, and cerebral hemorrhage incidence between the two groups. In addition, the incidence of bradycardia, hypotension, and the percentage of patients requiring intervention for these adverse events did not differ significantly between groups. In addition, the incidence of tachycardia (p=0·032), hypertension (p=0·026), and hypoxemia (p=0·027) were significantly lower in the Dex group than in the placebo group. Correspondingly, the proportion of patients requiring intervention for hypertension (p=0·025) and hypoxemia (p=0·034) was lower than those in the placebo group. The percentage of patients in the Dex group who changed the mode of infusion (including reducing the infusion rate or temporarily or permanently interrupting the infusion) was significantly higher (p=0·038) compared to the placebo group (Table 3).

**Table 3.**
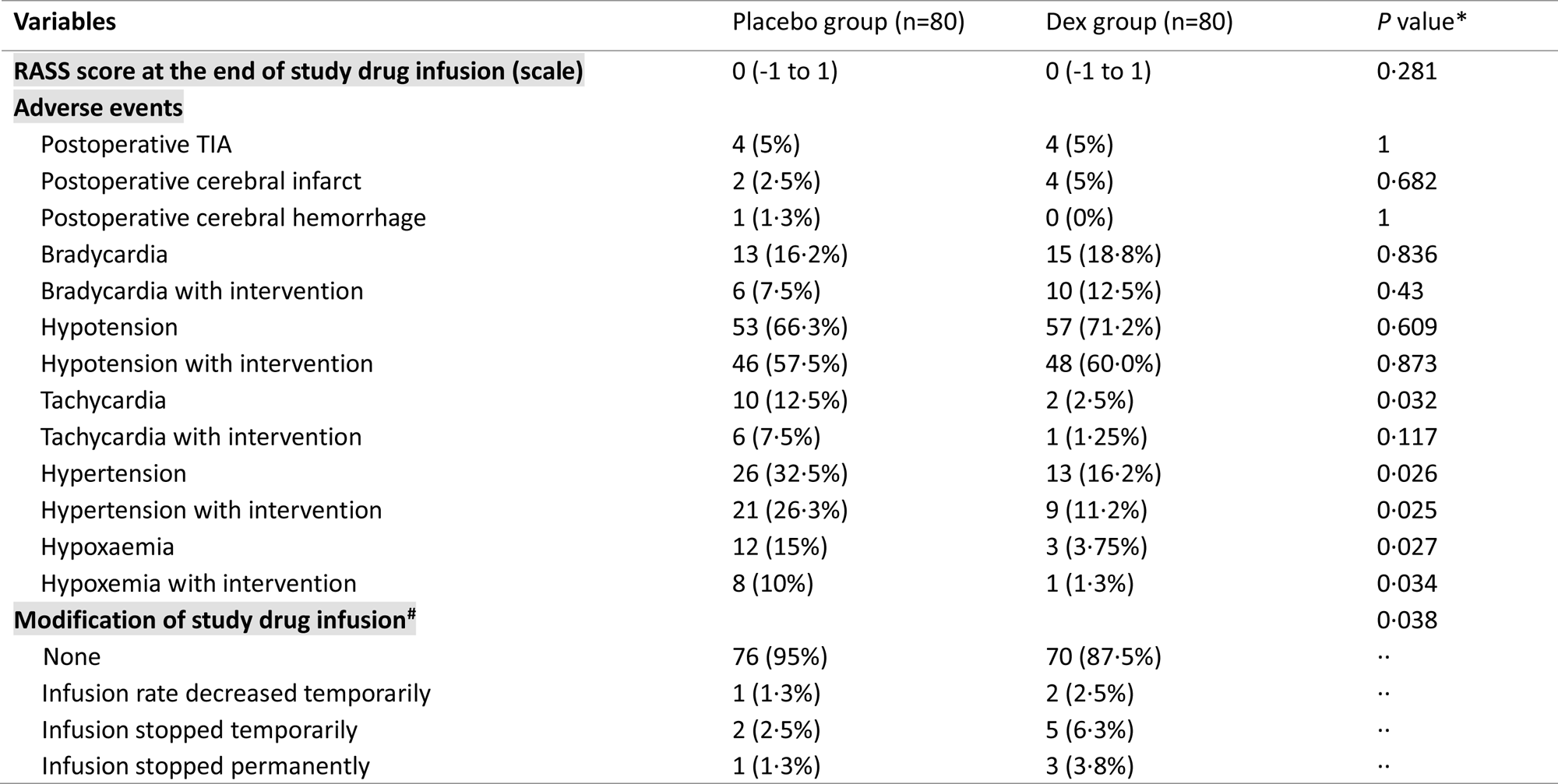
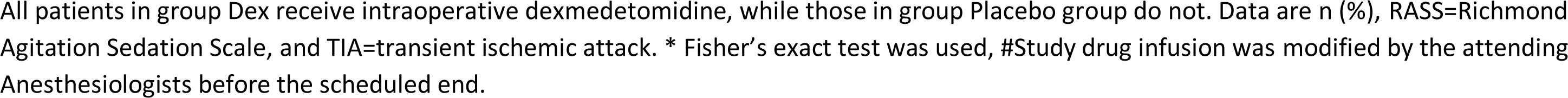
Adverse events.

### Metabolites

According to the score plot of the supervised orthogonal partial least squares discriminant analysis (OPLS-DA) model (Figures S4 A and B), the post-operative 6h and pre-anesthesia data were remarkably separated.

For the placebo group, the hierarchical clustering data showed the differences in the expression levels among 13 different metabolites (Figure 2A). In the placebo group, eight molecules were downregulated, and five were upregulated after CAS surgery at 6h compared to pre-anesthesia levels. For example, the expression levels of TAGs were significantly decreased in the Post-operative 6h group, especially the TAG (54:6) and TAG (54:7). In contrast, the expression levels of cholersteryl ester (CE) and Sphingomyelin (SM) were increased after CAS surgery. For the Dex group, the hierarchical clustering of 18 differential metabolites was found (Figure 2B). There were nine metabolites downregulated and upregulated after surgery, respectively. The TAGs were significantly downregulated post-operatively in the Dex group, especially TAG (54:5) and TAG (54:8). In addition, the CE and SM species were higher after surgery. As a result, LPE lipids were significantly increased after surgery in the Dex group compared to the placebo group.

**Figure 2.**
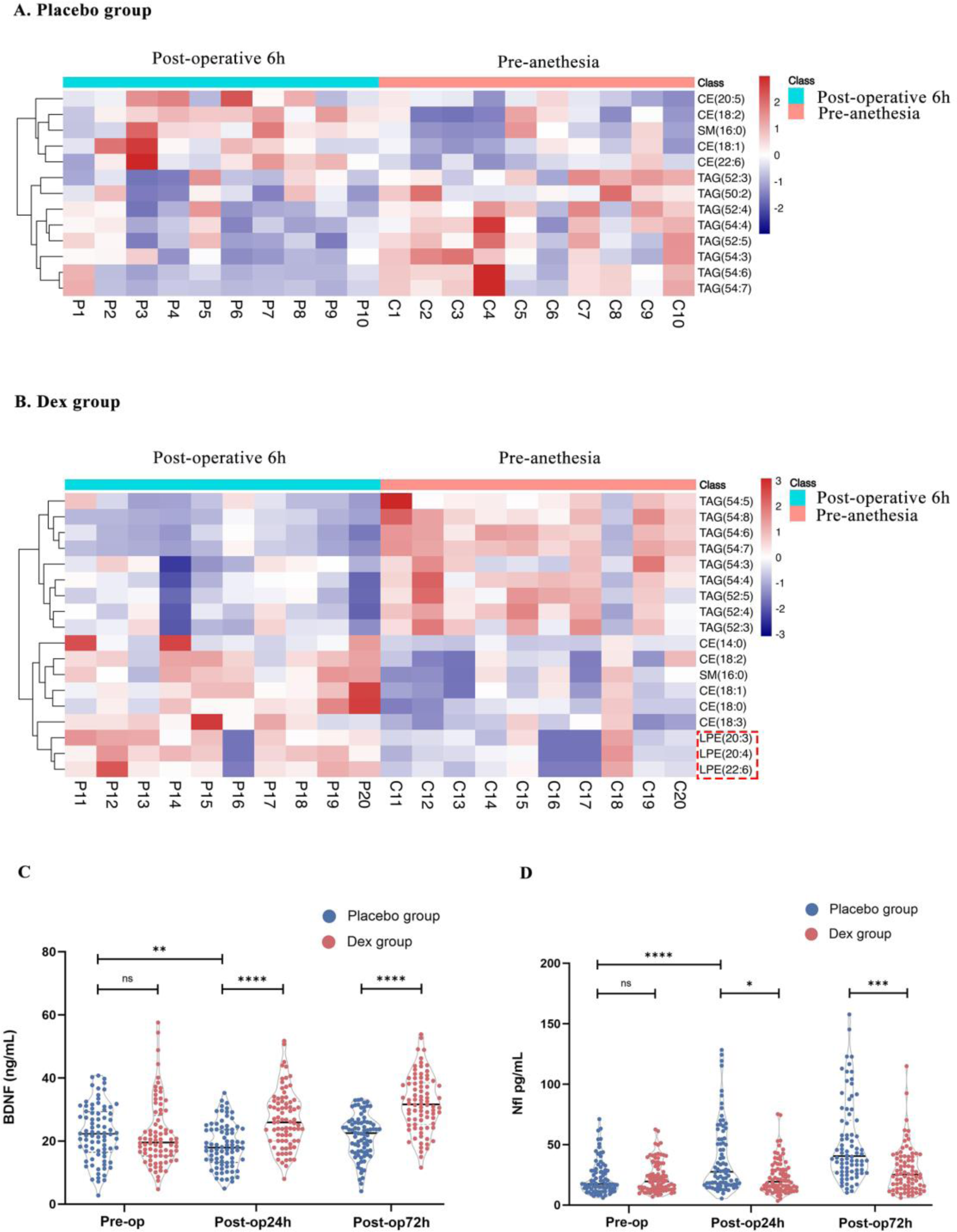
Significant Differential of Metabolites and serum BDNF and Nfl for pre-anesthesia and post-operative. (A) The differential metabolites of the Placebo group pre-anesthesia (C1-C10) and post-operative 6h (P1-P10) were compared, and the hierarchical clustering result was shown in a heat map. The X-axis represents sample names, and the Y-axis represents differential metabolites. The color blue to red indicates the expression abundance of metabolites from low to high. Bule: down-regulated, red up-regulated. (B) The differential metabolites of the Dex group pre-anesthesia (C11-C20) and post-operative 6h (P11-P20) were compared, and the hierarchical clustering result was shown in the heat map. Bule: down-regulated, red up-regulated. (C) Serum brain-derived neurotrophic factor concentration (BDNF) before (Pre-op) and 24h, 72h after the surgery (Post-op24h, Post-op72h) of two groups (n = 80). ns: P > 0·05. **:P ≤ 0·01. ****:P ≤ 0·0001. (D) Serum Neurofilament Light (Nfl) before (Pre-op) and 24h, 72h after the surgery (Post-op24h, Post-op72h) of two groups (n = 80). ns: P > 0·05. *: P < 0·05. ***: P < 0·001. ****: P < 0·0001.

The correlation analysis based on 13 differential metabolites was illustrated in Figure S4C. TAG lipids were strongly and positively correlated with their subtypes, except TAG (50:2) showed a weak correlation and negatively correlated with other CE and SM lipids. In addition, CE and SM lipid metabolites showed a positive correlation. The correlation analysis data for the Dex group was established in Figure S4D, whereby TAG lipids were strongly and positively correlated with their subtypes and with CE, SM, and LPE lipids. The significant Differential Metabolites for the whole dataset are presented in Figure S5.

### Biomarkers

Serum brain-derived neurotrophic factor (BDNF) was significantly increased (p<0·0001) in the Dex group at the post-operative 24 and 72 hr relative to the pre-operation (Figure 2C). These were significantly higher in the Dex group than in the placebo group at postoperative 24 and 72 hr (Figure 2C), respectively. Serum neurofilament light chain (Nfl) was significantly increased (p<0·0001) in the placebo group at post-operative 24 and 72 hr relative to the pre-operation. However, the Nfl was significantly lower (p<0·05, p<0·01) in the Dex group than in the placebo group at post-operative 24 and 72 hr, respectively (Figure 2D).

## DISCUSSION

In this randomized study, we showed that low-dose dexmedetomidine administered for the initial three days after carotid artery stenting (CAS) surgery significantly decreased the occurrence of cerebral hyperperfusion (CH) and its associated syndrome (CHS). However, there was a noteworthy improvement in the subjectively perceived sleep quality and mitigation of pain at rest, together with a shorter hospital stay and a lower frequency of adverse events, including hypertension and hypoxemia. Intriguingly, the administration of dexmedetomidine led to upregulation of BDNF, downregulation of Nfl, and upsurge of LPE (22:6) expression levels when compared to the placebo group. Notwithstanding, the intervention failed to significantly reduce the National Institute of Health Stroke Scale (NIHSS) and Modified Rankin Scale (mRS).

CHS is thought to be associated with multiple factors, including impaired brain autoregulation, blood-brain barrier destruction, nitric oxide and oxygen-free radical dis-regulation, baroreceptor dysfunction, and trigeminal neurovascular reflex disorder. In addition, the compromised ability to regulate the fraction of oxygen uptake in ischemic brain tissue was also considered to be a pathogenic CHS factor.^15^ The updated meta-analyses and pertinent clinical trials in Carotid Artery Stenosis (CAS) were reviewed comprehensively. Among them, Moulakakis et al.,^16^ encompassing nine studies and 4,446 patients, reported the incidence of CHS to be 1·16% (range 0·44-11·7%), while Garcia et al. ^17^ assessed 757 patients and reported 2·9% (22) cases of CHS. These incidences imply a lower prevalence than the placebo group (13·75%) in our study. This disparity may be due to our patients with severe internal carotid artery stenosis and complicated conditions who, therefore, received surgery under general anesthesia.

Ullery et al.,^18^ retrospectively reviewed 257 CAS surgeries and found that 63% of such procedures produced clinically significant hemodynamic instability. Hemodynamic instability is prevalent after CAS and also constitutes to be an independent risk factor for CHS. In this trial, patients were not given with a dexmedetomidine loading dose but only with a sub-sedative infusion rate (0·1 μg/kg/h). Therefore, Dex administration made stable hemodynamics and its adverse effects of hypotension and bradycardia were hardly seen (Table 3). In addition, no significant variations in the RASS scores between the two groups were observed, attesting to the absence of notable sedation triggered by low-dose dexmedetomidine, a result that mirrors previous studies.^5^ The present study also suggested that Dex might reduce the incidence of CHS due to decreasing rest-pain (Table 2), considering that severe headaches and subsequent increasing blood pressure may worsen hyperperfusion of brain tissue, intensifying pain and potentially resulting in severe brain oedema, cerebral hemorrhage, and death. A meta-analysis demonstrated that highly increased CBF might predict CHS,^4^ with 47% of affected individuals developing hemorrhagic stroke, while 54% of CHS cases were either dead or to be permanent disable. Our data also indicated that Dex infusion notably enhanced subjective sleep quality in the post-operative period. Dex stimulates pre- and post-synaptic alpha-2 receptors in the ventricular brain regions, inducing hypnosis akin to natural sleep, providing patients with the ability to be readily awakened from an unconscious state while remaining cooperative.^19^

Furthermore, in addition to improvements in hypoxemia, analgesia and sleep, the reduction of CH and CHS elicited by dexmedetomidine may also be attributed to its direct neuroprotective and cytoprotective effects.^20, 21^ A previous randomized control trial conducted in an intensive care setting demonstrated that dexmedetomidine attenuated delirium incidences in geriatric patients.^5^ Correspondingly, this study exhibits that Dexmedetomidine significantly upregulated neuroprotective factor brain-derived neurotrophic factor (BDNF) and downregulated neuronal injury biomarker neurofilament light chain (Nfl) in serum. Degos et al.,^22^ posited that Dexmedetomidine increased BDNF expression in astrocytes through an extracellular signal-regulated kinase-dependent pathway, whilst the potent neuroprotection and multiple neurological modulations of BDNF have been well documented.^23^

Lipids in the brain consist primarily of cholesterol (30%) and glycerophospholipids (49%)^15, 24^ Following a stroke and subsequent disruption of the blood-brain barrier, metabolites of these brain lipids enter the peripheral bloodstream and can be detected therein. Numerous studies demonstrated the role of lipid metabolites in regulating cellular homeostasis, and lipid metabolism dysregulation is the pathogenesis and also indicates the progression of a range of human diseases, including diabetes and neurodegeneration.^25^ To our best knowledge, this is the first study to obtain lipidomics profiling and examine its metabolite composition before and after undergoing CAS surgery. In this study, the LPE lipids, particularly the LPE (22:6 in the Dex group), were increased significantly, which was absent in the Placebo group following CAS surgery. Moreover, the TAG was considerably downregulated, specifically in relation to TAG (54:5) and TAG (54:8), whereas CE and SM levels were increased after CAS surgery in both the Placebo and Dex groups. The whole dataset analysis revealed that the top three signaling pathways enriched were cholesterol metabolism, fat digestion and absorption, and ovarian steroidogenesis. Previous studies showed a positive correlation between triglyceride and stroke and suggested implementing a strategy to lower triglyceride levels to reduce stroke risk.^26^ Another multi-center clinical study reported that intracranial artery stenosis patients had high triacylglycerol, a significant risk factor for ischemic stroke.^27^ In line with the previous study,^28^ Dex treatment decreased triacylglycerol levels. LPE, a type of phospholipid (PLs), plays a role in neurotransmission within the hippocampus and potentially functions as a biomarker for ischemic stroke. Mice suffering from global cerebral ischemia exhibited decreased levels of LPE (LPE 18:1, 20:3, and 22:6), and the long-term cognitive impairment was associated with a decline of hippocampal phospholipids.^29^ In line with these, we found that Dex treatment increased LPE levels (notably, 20:4 and 18:0).^30^ All these suggested that Dex use protects against the potential injurious risk to the brain.

### Limitations

Our study has limitations. Firstly, it is a single center, and whether the strategy reported herein can be applied widely is yet to be studied further. Secondly, this study only included patients who underwent CAS under general anesthesia due to complicated conditions. However, extracranial carotid artery stenosis is typically performed under local anesthesia; therefore, whether our strategy can be applied to those patients is still yet to be discovered. Thirdly, although the trial design and screening process were rigorously randomized, certain patients displayed imbalanced baseline and perioperative parameters between the two groups. However, this is not uncommon in randomized controlled trials. Fourthly, in our study, the onset of CHS was monitored for the first three days after surgery, whilst the incidence of CHS can occur within the first seven days post-surgery.^13^ Therefore, the ‘long term’ effect of Dex in preventing CH and CHS is also unknown and awaits further investigation.

### Conclusions

In summary, our study suggested that low dose dexmedetomidine significantly reduced the incidence of cerebral hyperperfusion syndrome and cerebral hyperperfusion in individuals undergoing carotid artery stenting. This was likely associated with hemodynamic stability and CBF reduction, decreasing brain metabolism, improving sleep quality and relieving pain to ultimately achieve neuroprotection. However, the strategy we reported here is subjected to further study before implementation into clinical practice.

## Author Contributions

Conceptualization: EC, TL, JZ and DM. Resources: JZ and DM. Data curation: MS, GC, LL. Formal Analysis: MS. Supervision: TL, JZ and DM. Funding acquisition: EC, JZ and DM. Validation: LW, LL. Investigation: HZ, MS. Methodology: EC, LW, TL, JZ and DM. Writing-original draft: EC, LW, JZ, TL, JZ and DM. Project administration: EC, LW, JZ, HZ, GC, LL. Writing-review & editing: EC, TL, JZ and DM.

## Conflict of Interest Disclosures

None reported.

## Funding/Support

National Natural Science Foundation of China (No. 82271288) and Henan Provincial Science and Technology Research Project (No. 202102310124).

## Data Availability

Will individual participant data be available (including data dictionaries)? Yes What data in particular will be shared? All of the individual participant data collected during the trial, after de-identification. What other documents will be available? Study protocol, statistical analysis plan, Informed consent form, clinical study report, analytic code When will data be available (start and end dates)? Following, publication With whom? Anyone who wishes to access the data For what types of analyses? Any purpose By what mechanism will data be made available? Data are available indefinitely at link to be included

## Acknowledgments

The authors gratefully acknowledge Prof. Ziliang Wang (Department of Cerebrovascular Disease, People’s Hospital of Zhengzhou University, Henan, China) for his help in cerebral hyperperfusion syndrome consultation. The authors also thank Prof. Liangfu Zhu (Department of Cerebrovascular Disease, People’s Hospital of Zhengzhou University, Henan, China) for his critical comments during manuscript preparation.

---

**Figure S1.**
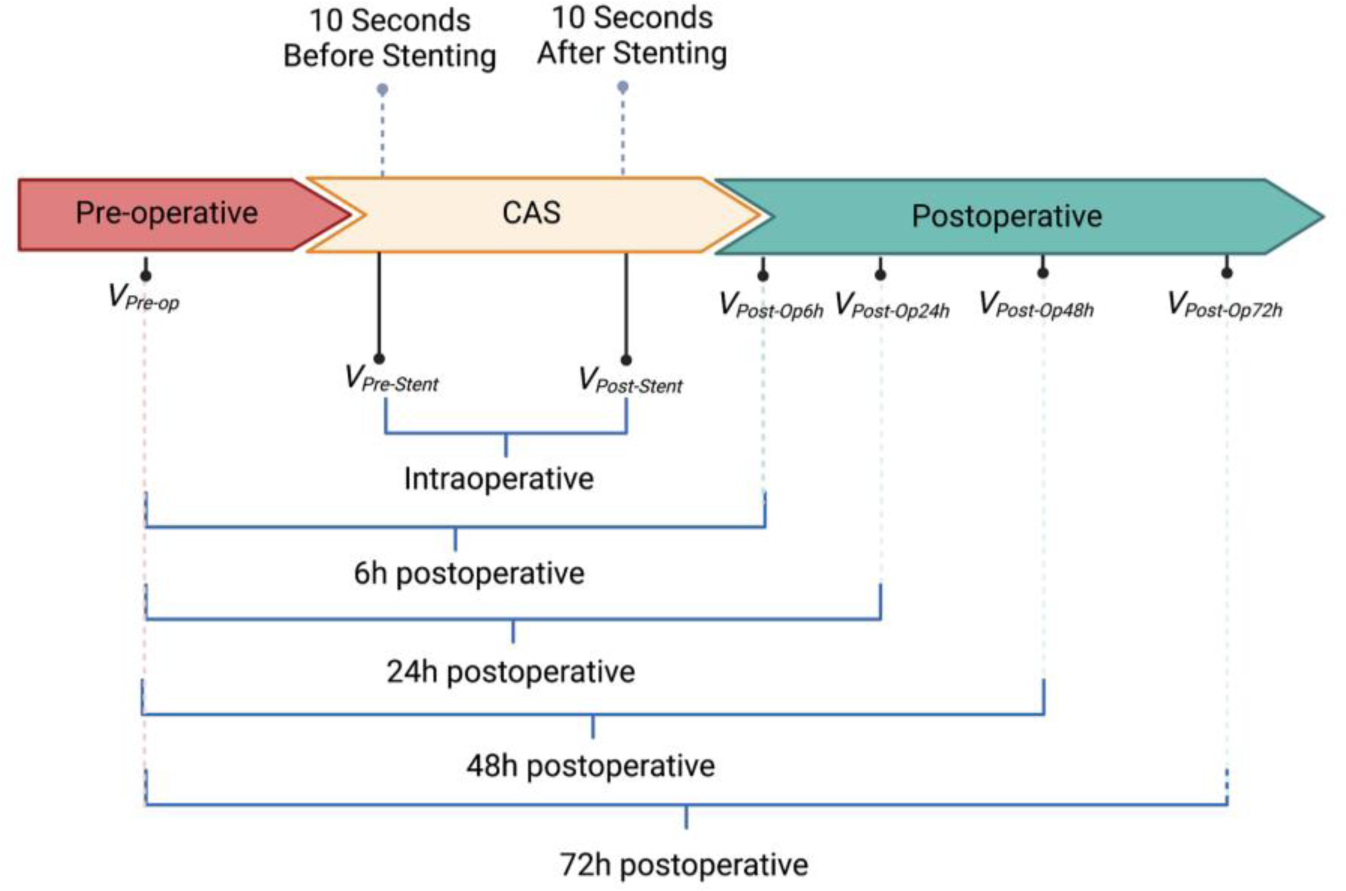
The transcranial doppler measurements’ timeframe. Transcranial doppler measurements (blue bracket) in patients undergoing carotid artery stenting (CAS) at different timeframes for prediction of cerebral hyperperfusion.

**Figure S2.**
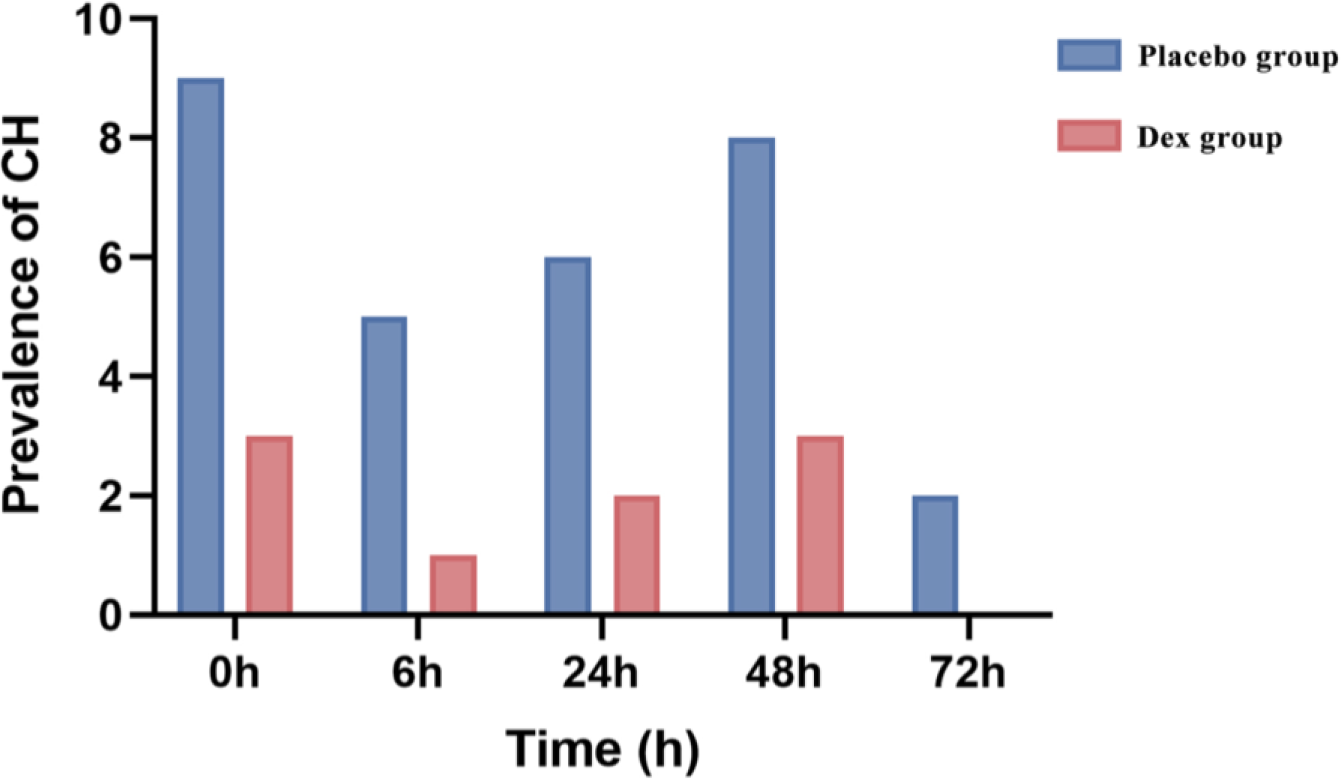
The incidence of cerebral hyperperfusion.

**Figure S3.**
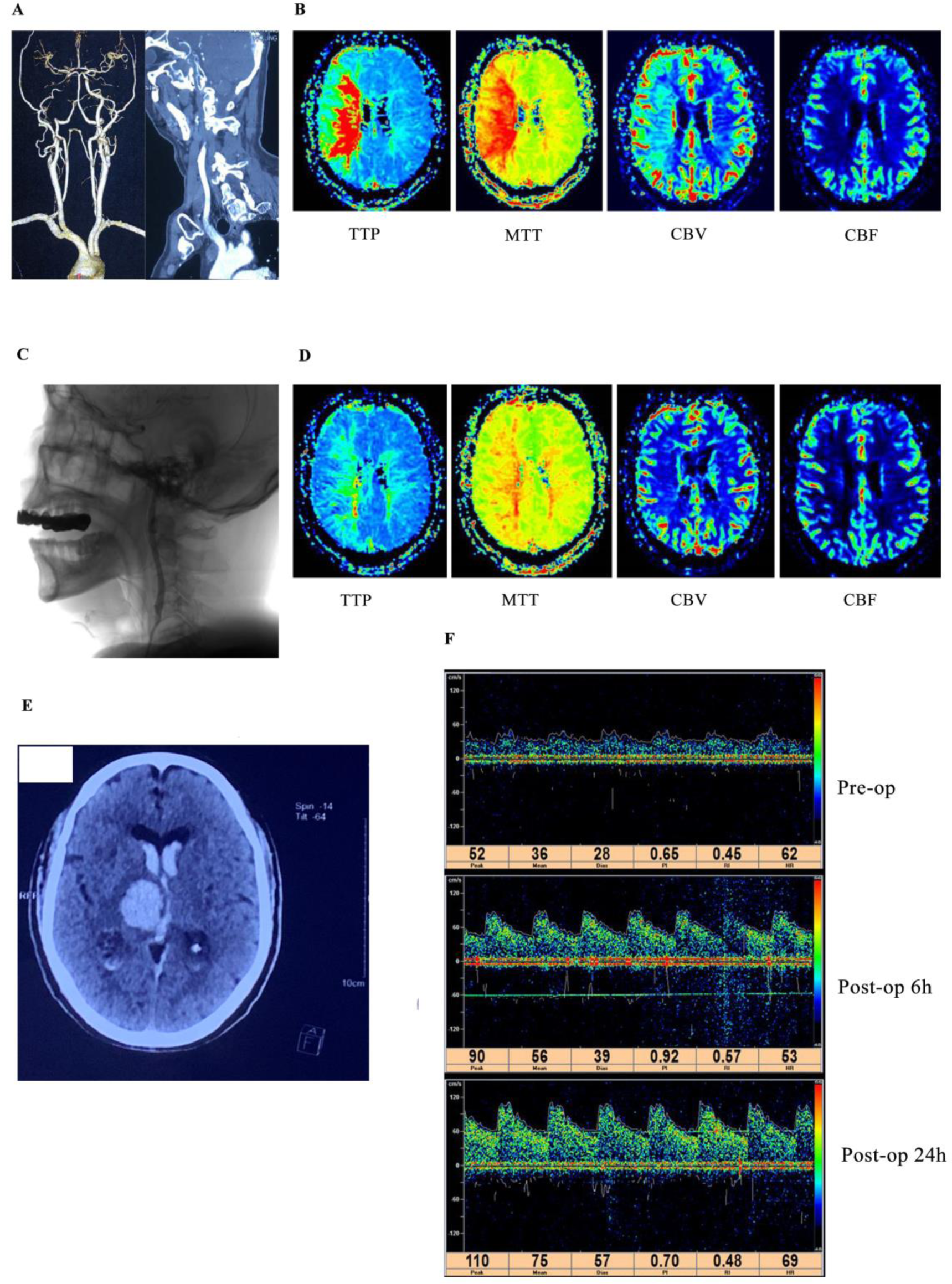
Data of death cases. (A). CTA (CT angiography): prolonged stage occlusion of the right internal carotid artery, and the distal vessels were normal. (B). PWI (Perfusion weighted imaging): delayed perfusion and insufficient compensation in the proper right middle cerebral artery territory. (C). Postoperative lateral radiography. (D). Postoperative PWI: Right cerebral perfusion was significantly improved. (E). Five days after surgery, the patient suddenly developed coma and dilated pupils. CT showed extensive hemorrhage in the ipsilateral cerebral hemisphere. (F). Transcranial doppler showed MCAVmean of middle cerebral artery Pre-op, Post-op6h, Post-op24h after operation were 36, 56, 75 cm/s, respectively. Hyperperfusion was diagnosed with a 100% increase in MCAVmean Post-op24h compared with that Pre-op. MCAVmean: mean blood flow velocity in the middle cerebral artery.

**Figure S4.**
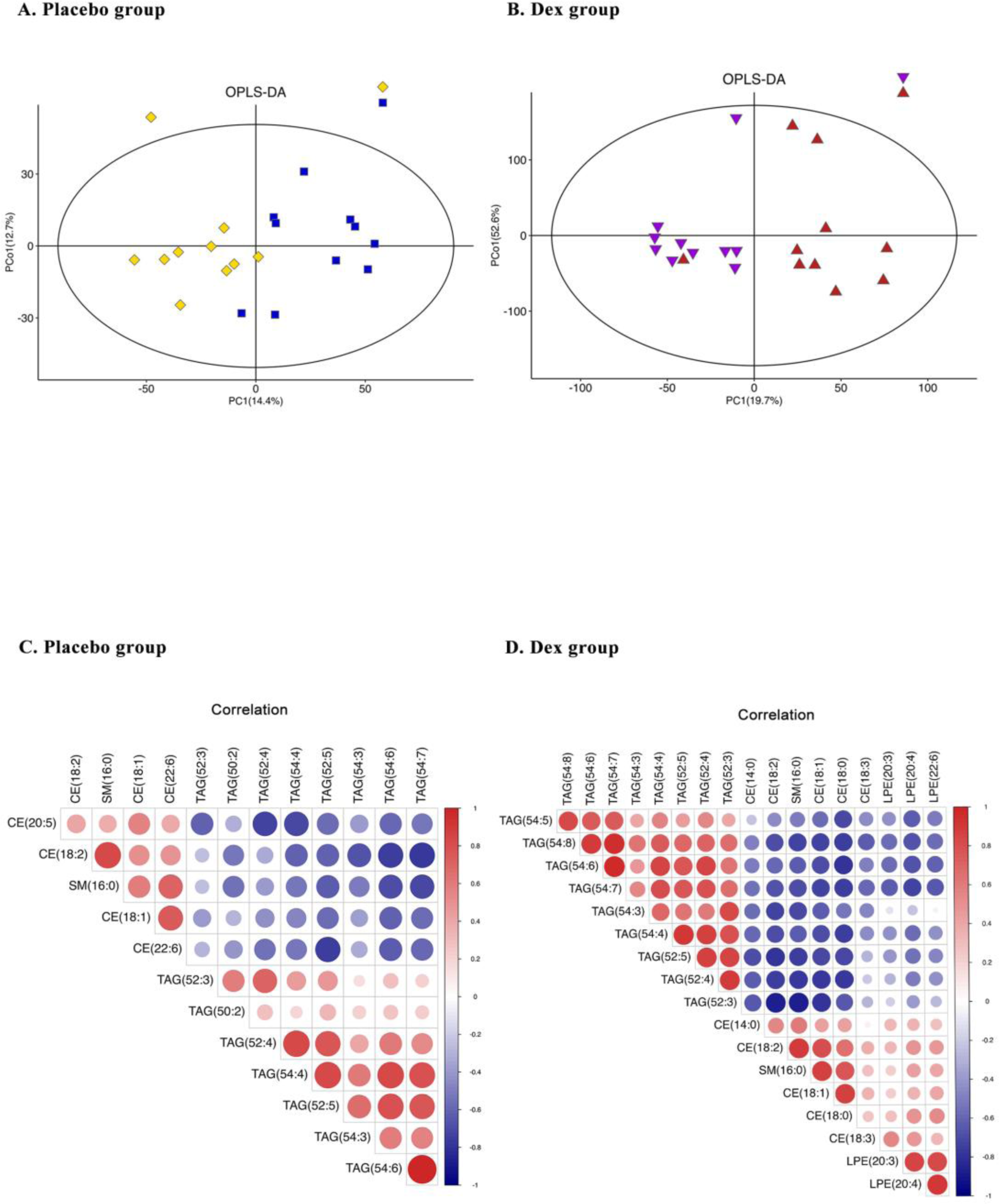
The OPLS-DA and correlation analysis of metabolites. (A) According to the score plot of the supervised orthogonal partial least squares discriminant analysis (OPLS-DA) model for the placebo group. (B) According to the score plot of the supervised orthogonal partial least squares discriminant analysis (OPLS-DA) model for the Dex group. (C) Correlation analysis diagram of 13 different metabolites from the Placebo group. The positive correlation is red, and the negative correlation is blue. Pearson correlation coefficient was used for correlation analysis. (D) Correlation analysis diagram of 18 different metabolites from the DEX group.

**Figure S5.**
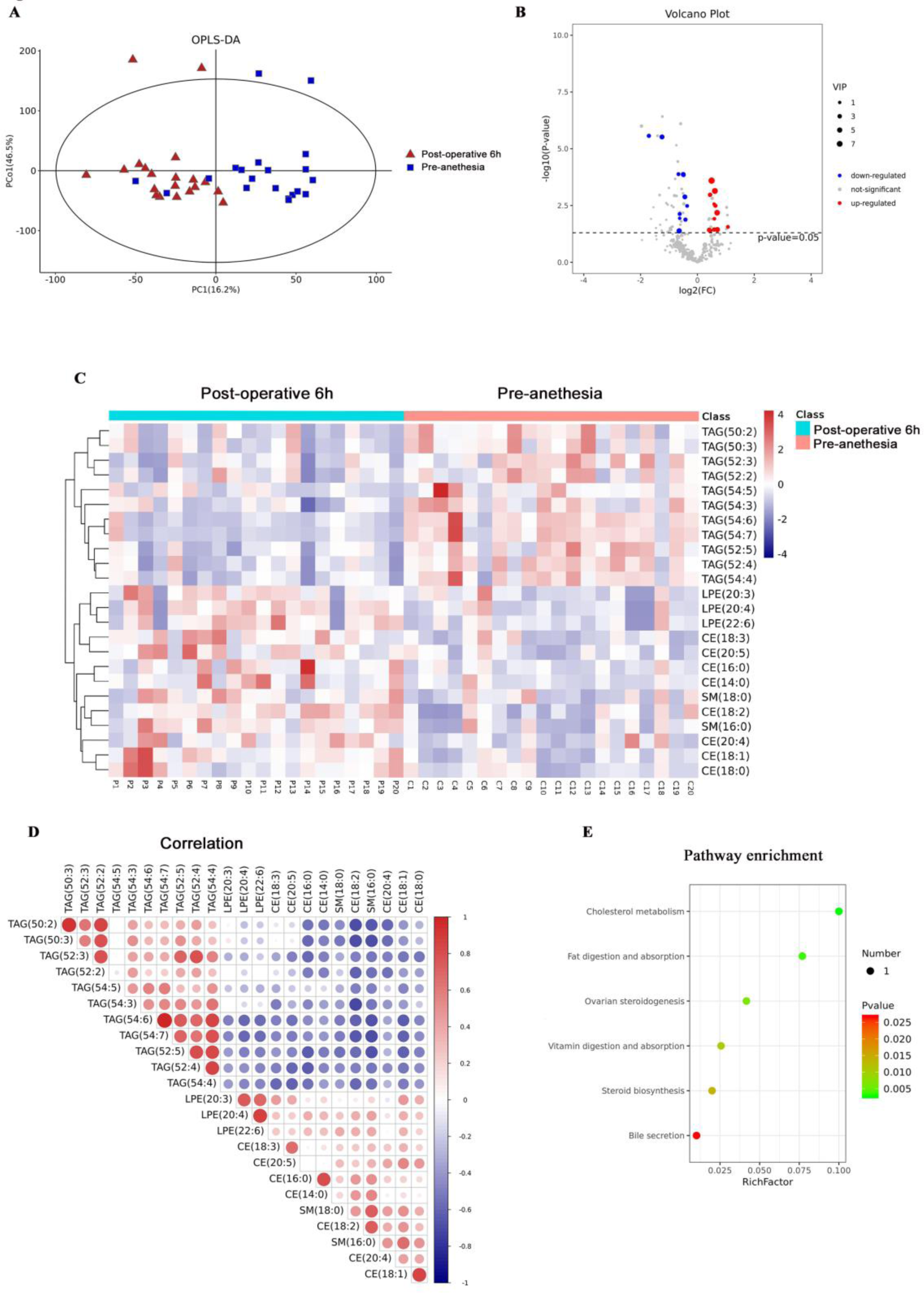
Significant Differential Metabolites for the whole dataset. (A). According to the score plot of the supervised orthogonal partial least squares discriminant analysis (OPLS-DA) model, the post-operative 6h group and the pre-anesthesia group were markedly separated, indicating that there was a significant change in metabolites between the two groups. At the same time, the effectiveness and analytical power of the model are proven. Red: post-operative 6h group, blue: pre-anesthesia group. (B). Student T-test and fold change analysis in univariate statistical analysis were used to compare the differential metabolites before and after CAS surgery. The results were visualized in the volcano diagram. Bule: down-regulated, red up-regulated. (C). The result of hierarchical clustering, which illustrates the differences in expression levels among these 24 different metabolites, was visualized in the heat map. 11 molecules had downregulated expression, and 13 had upregulated expression levels after CAS surgery. Compared with the pre-anesthesia group, the expression levels of triglyceride and triacylglycerol (TAG) were significantly lower in the after CAS group, especially the expression of TAG (50:2). Still, the expression levels of Lysphosphatidylethanolamine (LPE), cholesteryl ester (CE), and Sphingomyelin (SM) were higher after CAS surgery. Bule: down-regulated, red up-regulated (D). The result of correlation analysis based on 24 differential metabolites was visualized. TAG lipids were strongly positively correlated with their subtypes, except for TAG (52:2) and TAG (54:5), and negatively associated with other LPE, CE and SM lipids. LPE, CE, and SM lipid metabolites showed a weak positive correlation. Bule: negatively correlated, red positively correlated. (E). With P-value < 0·05 as the threshold, the significantly enriched signal pathway and the corresponding enrichment of differential metabolites in these pathways. The top three signaling pathways with significant accumulation were cholesterol metabolism, fat digestion and absorption and ovarian steroidogenesis signaling pathways, P <0·01.

## Notes

### Competing Interest Statement

The authors have declared no competing interest.

### Clinical Trial

ChiCTR1900024416

### Author Declarations

The Ethics Committee of People's Hospital of Zhengzhou University (2019-43), China

## Reference

1. Collaborators GBDS. Global, regional, and national burden of stroke and its risk factors, 1990-2019: a systematic analysis for the Global Burden of Disease Study 2019. Lancet Neurol. 2021;20:795–820. doi: 10.1016/S1474-4422(21)00252-0

2. Sandset EC, Anderson CS, Bath PM, Christensen H, Fischer U, Gasecki D, Lal A, Manning LS, Sacco S, Steiner T, et al. European Stroke Organisation (ESO) guidelines on blood pressure management in acute ischaemic stroke and intracerebral haemorrhage. Eur Stroke J. 2021;6:II. doi: 10.1177/23969873211026998

3. van Mook WN, Rennenberg RJ, Schurink GW, van Oostenbrugge RJ, Mess WH, Hofman PA, de Leeuw PW. Cerebral hyperperfusion syndrome. Lancet Neurol. 2005;4:877–888. doi: 10.1016/s1474-4422(05)70251-9

4. Huibers AE, Westerink J, de Vries EE, Hoskam A, den Ruijter HM, Moll FL, de Borst GJ. Editor’s Choice - Cerebral Hyperperfusion Syndrome After Carotid Artery Stenting: A Systematic Review and Meta-analysis. Eur J Vasc Endovasc Surg. 2018;56:322–333. doi: 10.1016/j.ejvs.2018.05.012

5. Su X, Meng ZT, Wu XH, Cui F, Li HL, Wang DX, Zhu X, Zhu SN, Maze M, Ma D. Dexmedetomidine for prevention of delirium in elderly patients after non-cardiac surgery: a randomised, double-blind, placebo-controlled trial. Lancet. 2016;388:1893–1902. doi: 10.1016/S0140-6736(16)30580-3

6. Karlsson BR, Forsman M, Roald OK, Heier MS, Steen PA. Effect of dexmedetomidine, a selective and potent alpha 2-agonist, on cerebral blood flow and oxygen consumption during halothane anesthesia in dogs. Anesth Analg. 1990;71:125–129. doi: 10.1213/00000539-199008000-00003

7. Alam A, Suen KC, Hana Z, Sanders RD, Maze M, Ma D. Neuroprotection and neurotoxicity in the developing brain: an update on the effects of dexmedetomidine and xenon. Neurotoxicol Teratol. 2017;60:102–116. doi: 10.1016/j.ntt.2017.01.001

8. Drummond JC, Dao AV, Roth DM, Cheng CR, Atwater BI, Minokadeh A, Pasco LC, Patel PM. Effect of dexmedetomidine on cerebral blood flow velocity, cerebral metabolic rate, and carbon dioxide response in normal humans. Anesthesiology. 2008;108:225–232. doi: 10.1097/01.anes.0000299576.00302.4c

9. Aalling NN, Nedergaard M, DiNuzzo M. Cerebral Metabolic Changes During Sleep. Curr Neurol Neurosci Rep. 2018;18:57. doi: 10.1007/s11910-018-0868-9

10. Chimowitz MI, Lynn MJ, Howlett-Smith H, Stern BJ, Hertzberg VS, Frankel MR, Levine SR, Chaturvedi S, Kasner SE, Benesch CG, et al. Comparison of warfarin and aspirin for symptomatic intracranial arterial stenosis. N Engl J Med. 2005;352:1305–1316. doi: 10.1056/NEJMoa043033

11. Pennekamp CW, Tromp SC, Ackerstaff RG, Bots ML, Immink RV, Spiering W, de Vries JP, Kappelle LJ, Moll FL, Buhre WF, et al. Prediction of cerebral hyperperfusion after carotid endarterectomy with transcranial Doppler. Eur J Vasc Endovasc Surg. 2012;43:371–376. doi: 10.1016/j.ejvs.2011.12.024

12. Fassaert LMM, Immink RV, van Vriesland DJ, de Vries JPM, Toorop RJ, Kappelle LJ, Westerink J, Tromp SC, de Borst GJ. Transcranial Doppler 24 Hours after Carotid Endarterectomy Accurately Identifies Patients Not at Risk of Cerebral Hyperperfusion Syndrome. Eur J Vasc Endovasc Surg. 2019;58:320–327. doi: 10.1016/j.ejvs.2019.04.033

13. Galyfos G, Sianou A, Filis K. Cerebral hyperperfusion syndrome and intracranial hemorrhage after carotid endarterectomy or carotid stenting: A meta-analysis. J Neurol Sci. 2017;381:74–82. doi: 10.1016/j.jns.2017.08.020

14. Sessler CN, Gosnell MS, Grap MJ, Brophy GM, O’Neal PV, Keane KA, Tesoro EP, Elswick RK. The Richmond Agitation-Sedation Scale: validity and reliability in adult intensive care unit patients. Am J Respir Crit Care Med. 2002;166:1338–1344. doi: 10.1164/rccm.2107138

15. Ng FC, Churilov L, Yassi N, Kleinig TJ, Thijs V, Wu TY, Shah DG, Dewey HM, Sharma G, Desmond PM, et al. Microvascular Dysfunction in Blood-Brain Barrier Disruption and Hypoperfusion Within the Infarct Posttreatment Are Associated With Cerebral Edema. Stroke. 2022;53:1597–1605. doi: 10.1161/STROKEAHA.121.036104

16. Moulakakis KG, Mylonas SN, Sfyroeras GS, Andrikopoulos V. Hyperperfusion syndrome after carotid revascularization. J Vasc Surg. 2009;49:1060–1068. doi: 10.1016/j.jvs.2008.11.026

17. Gonzalez Garcia A, Moniche F, Escudero-Martinez I, Mancha F, Tomasello A, Ribo M, Delgado-Acosta F, Ochoa JJ, de Las Heras JA, Lopez-Mesonero L, et al. Clinical Predictors of Hyperperfusion Syndrome Following Carotid Stenting: Results From a National Prospective Multicenter Study. JACC Cardiovasc Interv. 2019;12:873–882. doi: 10.1016/j.jcin.2019.01.247

18. Ullery BW, Nathan DP, Shang EK, Wang GJ, Jackson BM, Murphy EH, Fairman RM, Woo EY. Incidence, predictors, and outcomes of hemodynamic instability following carotid angioplasty and stenting. J Vasc Surg. 2013;58:917–925. doi: 10.1016/j.jvs.2012.10.141

19. Alexopoulou C, Kondili E, Diamantaki E, Psarologakis C, Kokkini S, Bolaki M, Georgopoulos D. Effects of dexmedetomidine on sleep quality in critically ill patients: a pilot study. Anesthesiology. 2014;121:801–807. doi: 10.1097/ALN.0000000000000361

20. Wang K, Wu M, Xu J, Wu C, Zhang B, Wang G, Ma D. Effects of dexmedetomidine on perioperative stress, inflammation, and immune function: systematic review and meta-analysis. Br J Anaesth. 2019;123:777–794. doi: 10.1016/j.bja.2019.07.027

21. Sun YB, Zhao H, Mu DL, Zhang W, Cui J, Wu L, Alam A, Wang DX, Ma D. Dexmedetomidine inhibits astrocyte pyroptosis and subsequently protects the brain in in vitro and in vivo models of sepsis. Cell Death Dis. 2019;10:167. doi: 10.1038/s41419-019-1416-5

22. Degos V, Charpentier TL, Chhor V, Brissaud O, Lebon S, Schwendimann L, Bednareck N, Passemard S, Mantz J, Gressens P. Neuroprotective effects of dexmedetomidine against glutamate agonist-induced neuronal cell death are related to increased astrocyte brain-derived neurotrophic factor expression. Anesthesiology. 2013;118:1123–1132. doi: 10.1097/ALN.0b013e318286cf36

23. Khatoon S, Samim M, Dahalia M, Nidhi. Fisetin provides neuroprotection in pentylenetetrazole-induced cognition impairment by upregulating CREB/BDNF. Eur J Pharmacol. 2023;944:175583. doi: 10.1016/j.ejphar.2023.175583

24. Martin M, Vermeiren S, Bostaille N, Eubelen M, Spitzer D, Vermeersch M, Profaci CP, Pozuelo E, Toussay X, Raman-Nair J, et al. Engineered Wnt ligands enable blood-brain barrier repair in neurological disorders. Science. 2022;375:eabm4459. doi: 10.1126/science.abm4459

25. Brown HA, Thomas PG, Lindsley CW. Targeting phospholipase D in cancer, infection and neurodegenerative disorders. Nat Rev Drug Discov. 2017;16:351–367. doi: 10.1038/nrd.2016.252

26. Rothwell PM, Algra A, Amarenco P. Medical treatment in acute and long-term secondary prevention after transient ischaemic attack and ischaemic stroke. Lancet. 2011;377:1681–1692. doi: 10.1016/S0140-6736(11)60516-3

27. Ganesan V, Prengler M, McShane MA, Wade AM, Kirkham FJ. Investigation of risk factors in children with arterial ischemic stroke. Ann Neurol. 2003;53:167–173. doi: 10.1002/ana.10423

28. Lv ZP, Peng YZ, Zhang BB, Fan H, Liu D, Guo YM. Glucose and lipid metabolism disorders in the chickens with dexamethasone-induced oxidative stress. J Anim Physiol Anim Nutr (Berl). 2018;102:e706–e717. doi: 10.1111/jpn.12823

29. Sabogal-Guaqueta AM, Posada-Duque R, Cortes NC, Arias-Londono JD, Cardona-Gomez GP. Changes in the hippocampal and peripheral phospholipid profiles are associated with neurodegeneration hallmarks in a long-term global cerebral ischemia model: Attenuation by Linalool. Neuropharmacology. 2018;135:555–571. doi: 10.1016/j.neuropharm.2018.04.015

30. Nummela AJ, Laaksonen LT, Laitio TT, Kallionpaa RE, Langsjo JW, Scheinin JM, Vahlberg TJ, Koskela HT, Aittomaki V, Valli KJ, et al. Effects of dexmedetomidine, propofol, sevoflurane and S-ketamine on the human metabolome: A randomised trial using nuclear magnetic resonance spectroscopy. Eur J Anaesthesiol. 2022;39:521–532. doi: 10.1097/EJA.0000000000001591

